# Complex multi-environment gene interactions linked to the interplay between polysubstance dependence and suicidal behaviors

**DOI:** 10.1101/2020.01.14.20017509

**Authors:** Renato Polimanti, Daniel F. Levey, Gita A. Pathak, Frank R. Wendt, Yaira Z. Nunez, Robert J. Ursano, Ronald C. Kessler, Henry R. Kranzler, Murray B. Stein, Joel Gelernter

**Affiliations:** Department of Psychiatry, Yale School of Medicine, Yale University, West Haven, CT; Veteran Affairs CT Healthcare System, West Haven, CT; Center for the Study of Traumatic Stress, Department of Psychiatry, Uniformed Services University of the Health Sciences, Bethesda, MD, USA; Department of Health Care Policy, Harvard Medical School, Boston MA; University of Pennsylvania Perelman School of Medicine, Philadelphia, PA; Crescenz Veterans Affairs Medical Center, Philadelphia, PA; Department of Psychiatry, School of Medicine, University of California, San Diego, La Jolla, CA; Psychiatry Service, Veterans Affairs San Diego Healthcare System, San Diego, CA; Departments of Genetics and Neuroscience, Yale University School of Medicine, New Haven, CT 06510, USA

**Keywords:** addiction, suicide, comorbidity, genome-wide analyses, gene-environment interactions, mental health, polysubstance abuse

## Abstract

**Background and Aims:** Substance dependence diagnoses (SDs) are important risk factors for suicidal behaviors. We investigated the associations of multiple SDs with different suicidal behaviors and tested how genetic background moderates these associations.

**Design:** Multivariate logistic regression to investigate the associations of SDs with suicidal behaviors; structured linear mixed model to study multivariate gene– environment interactions.

**Setting:** The Yale-Penn cohort was recruited to investigate the genetics of SDs. The Army STARRS (Study to Assess Risk and Resilience in Servicemembers) cohort was recruited to evaluate mental health risk and resilience for suicidal behaviors among Army personnel.

**Participants:** Yale-Penn participants (N=15,557) were assessed via the Semi-Structured Assessment for Drug Dependence and Alcoholism. Army STARRS participants (N=11,236) were evaluated using the self-administered Composite International Diagnostic Interview Screening Scales.

**Measurements:** Lifetime self-reported suicidal behaviors (ideation, SI; planning; attempt, SA); Lifetime DSM-IV diagnoses and criteria for dependence on alcohol, cannabis, cocaine (CoD), opioid (OD), and nicotine (ND) (Yale-Penn); substance use disorder (SUD) (Army STARRS).

**Findings:** In Yale-Penn, lifetime polysubstance dependence was strongly associated with lifetime suicidal behaviors: individuals with five SDs showed increased odds ranging from OR=6.77 (95%CI=5.74-7.99) for SI to OR=3.61 (95%CI=2.7-4.86) for SA. In Army STARRS, SUD was associated with increased odds ranging from OR=2.88 (95%CI=2.6-3.19) for SI to OR=3.92 (95%CI=3.19-4.81) for SA. In Yale-Penn, we identified multivariate gene–environment interactions (Bayes factors, BF > 0) of SI with respect to a gene cluster on chromosome 16 (*LCAT*, p=1.82×10^−7^; *TSNAXIP1*, p=2.13×10^−7^; *CENPT*, p=2.32×10^−7^; *PARD6A*, p=5.57×10^−7^) for OD (BF=12.2), CoD (BF=12.1), ND (BF=9.2), and polysubstance dependence (BF=2.1).

**Conclusions:** Comorbidity of multiple SDs is a significant suicide risk factor and heritability of suicidal behaviors is partially moderated by multivariate gene interactions.

## Introduction

Individuals with substance dependence diagnoses (SDs) are a population with the high suicide risk. Compared to the general population, people with SDs are 10 to 14 times as likely to die by suicide and poly-drug abusers have 17-fold increase risk of suicide rates (1). Forty percent of patients seeking treatment for a SD report a history of suicide attempts (2); 56% of individuals who died by suicide in a U.S. psychological autopsy study had alcohol dependence (3); and of a sample of patients in substance abuse treatment, 39% percent of opiate-dependent and 43.5% of cocaine-dependent had attempted suicide (2, 4).

While there is a strong correlation between suicide-related behaviors and SDs, twin, family and adoption studies suggest that the genetic component of suicidal behavior (heritability ranging between 30-50%) is independent from SD (5). This was confirmed by polygenic risk score (PRS) analyses (6), consistent with a partially independent genetic diathesis to act on suicidal thoughts. SDs may thus interact with the genetic predisposition to suicidal behaviors. Under this scenario, a genome-wide gene-by-SD analysis may be a powerful tool to dissect the genetics of suicide. Most genetic studies of suicidal behaviors are candidate gene analyses that, as demonstrated for other psychiatric traits (7, 8), have limited power to detect the key molecular processes of suicide. Recently, several genome-wide association studies (GWAS) of suicidal behaviors have been conducted in large cohorts, identifying several risk loci and confirming a partial genetic overlap with depression (9-13). This supports the notion that genome-wide investigations are useful for investigating the predisposition to suicidal behaviors. However, to our knowledge, no gene-by-environment genome-wide interaction studies (GEWIS) of suicidal behaviors have yet been conducted. We previously showed that this approach is useful to identify the complex interactive mechanisms linking genetic background with the interplay between SDs and psychiatric and behavioral phenotypes (14-17). In particular, polysubstance dependence could potentially interact with the genetic background to influence suicidal behaviors. Accordingly, to test complex interactions related to the effect of multiple SDs, we applied a recently-developed linear mixed-model approach (18) that can efficiently test and characterize loci that interact with one or more environments, in this case diagnoses of substance use/abuse of various types.

The aim of this study was to investigate the association of multiple SDs (testing for each SD the individual effect independent from the other SDs; and the cumulative effect of having received multiple SD diagnoses) to several suicidal behaviors (i.e., ideation SI; planning, SP; and attempt, SA) and to test whether SDs interact with the individual genetic variability to increase SI risk. The analyses were conducted in two large cohorts (total sample size >26,000 individuals) representative of groups with different baseline suicide risk: substance abusers and military personnel.

## Methods

### Study Populations

Yale-Penn participants were recruited for studies of the genetics of drug or alcohol dependence in five eastern U.S. centers as described elsewhere (19-23). Subjects gave written informed consent as approved by the institutional review board at each site, and certificates of confidentiality were obtained from NIDA and NIAAA. Subjects were evaluated with the Semi-Structured Assessment for Drug Dependence and Alcoholism (SSADDA) (24) to derive DSM-IV lifetime SD diagnoses and other major psychiatric traits. In the present study, we used information regarding DSM-IV SD diagnoses and criterion counts related to alcohol dependence (AD), cannabis dependence (CaD), cocaine dependence (CoD), nicotine dependence (ND), and opioid dependence (OD). Detailed information regarding these phenotypic definitions is provided in our previous studies (19-23). Data regarding suicidal behaviors were derived from SSADDA items: SI “*Have you ever thought about killing yourself?”* ; persistent SI “*Did those thoughts persist for at least 7 days in a row?*” ; SP “*Did you have a plan?*” ; and SA “*Have you ever tried to kill yourself?*”. This phenotypic information was available for 15,557 participants. Full genome-wide data were available for a subset of these individuals via genotyping done with the Illumina HumanOmni1-Quad microarray, the Illumina HumanCoreExome array, or the Illumina Multi-Ethnic Global Array. Principal component (PC) analysis was conducted based on each genotyping array and for each ancestry group (African and European ancestries) separately using Eigensoft and SNPs that were common to the GWAS data sets and 1000 Genomes Phase 1 panel. Detailed information about the pre-imputation quality control pipeline is available in our previous studies (19-23). Data were imputed using Minimac3 implemented in the Michigan Imputation Server with the 1000 Genomes Phase 3 reference panel. Dosage data were transformed into best-estimate genotypes using PLINK2 (25), considering variants with info score ≥80% and minor allele frequency ≥1%. The present study only considered information regarding unrelated subjects. Related subjects were identified using genetic information as previously described (26) and, within each family group determined from genetic data, a subject was selected prioritizing retention of participants who reported the most extreme suicidal behaviors among those genetically related. The final sample for Yale-Penn sample in the present genome-wide analysis included 4,044 African-American and 3,407 European-American participants.

The Army STARRS (Study to Assess Risk and Resilience in Servicemembers) participants included individuals recruited from two different groups: The New Soldier Study (soldiers recruited at the start of their basic training at one of three Army installations); and The Pre-Post Deployment Study (a multiple-wave panel survey that collected baseline data from US Army soldiers in three brigade combat teams prior to their deployment to Afghanistan). All subjects gave written informed consent to participate. These procedures were approved by the Human Subjects Committees of all collaborating institutions. Detailed information about the design and conduct of the Army STARRS is available in a previous report (27). Every individual was diagnosed using a self-administered questionnaire, which included the adapted versions of the Composite International Diagnostic Interview Screening Scales (CIDI-SC) (28). As previously described (29), the CIDI-SC assessment was used to determine lifetime prevalence of 12 common lifetime DSM-IV mental disorders, including substance use disorder for alcohol and/or drugs combined (i.e., SUD_combined_). Suicidal behaviors were assessed using a modified version of the Columbia–Suicide Severity Rating Scale (30), which assesses the lifetime occurrence of SI (“*Did you ever in your life have thoughts of killing yourself*?” or “*Did you ever wish you were dead or would go to sleep and never wake up?*”), SP (“*Did you ever have any intention to act [on these thoughts/on that wish*]?” and, if so, “*Did you ever think about how you might kill yourself [e.g*., *taking pills, shooting yourself] or work out a plan of how to kill yourself?*”), and SA (“*Did you ever make a suicide attempt [i.e*., *purposefully hurt yourself with at least some intention to die]?*”). Army STARRS participants were genotyped using the Illumina OmniExpress and Exome array or the Illumina PsychChip array. Methods for genotyping, imputation, ancestry assignment and PC analysis were described previously (31). In the present study, we investigated 11,235 Army STARRS participants of European descent.

Table 1 reports the characteristics of the Yale-Penn and Army STARRS participants investigated in the present study.

**Table 1:** Characteristics of the Yale-Penn and Army STARRS participants investigated in the present study.

| **Yale-Penn, n=15,557** |  |
| --- | --- |
| Age, mean (SD) | 40 (11.8) |
| Sex, Women (%) | 7,187 (46) |
| Self-reported Population Group, n (%) |  |
| *Native American/American Indian* | 1,327 (9) |
| *Asian* | 101 (1) |
| *Pacific Islander* | 20 (<1) |
| *African-American/Black, not of Hispanic origin* | 6,027 (39) |
| *African-American/Black, of Hispanic origin* | 350 (2) |
| *Caucasian/White, not of Hispanic origin* | 6,060 (39) |
| *Caucasian/White, of Hispanic origin* | 881 (5) |
| DSM-IV diagnosis, n (%) |  |
| *Alcohol Dependence* | 7,481 (48) |
| *Cannabis Dependence* | 3,897 (25) |
| *Cocaine Dependence* | 8,662 (56) |
| *Nicotine Dependence* | 8,219 (52) |
| *Opioid Dependence* | 4,379 (28) |
| Polysubstance Dependence, n (%) |  |
| *One DSM-IV SD diagnosis* | 2,023 (13) |
| *Two DSM-IV SD diagnoses* | 2,942 (22) |
| *Three DSM-IV SD diagnoses* | 3,345 (22) |
| *Four DSM-IV SD diagnoses* | 2,419 (16) |
| *Five DSM-IV SD diagnoses* | 1,004 (6) |
| Suicidal behaviors, n (%) |  |
| *Ideation* | 6,112 (39) |
| *Persistent Ideation* | 1,450 (9) |
| *Planning* | 2,491 (16) |
| *Attempt* | 1,965 (13) |
| **Army STARRS, n=11,235** |  |
| Age, mean (SD) | 21 (5.2) |
| Sex, Women (%) | 1,163 (10) |
| SUD<sub>combined</sub>, n (%) | 2,848 (22) |
| Suicidal behaviors, n (%) |  |
| *Ideation* | 2,299 (20) |
| *Planning* | 446 (4) |
| *Attempt* | 389 (3) |

*Data Analysis*

The phenotypic association of SDs with suicidal behaviors (i.e., SI, SP, and SA) was calculated using multivariate logistic regression models via the computing environment R (32). In the Yale-Penn cohort, this analysis was conducted in the full sample (N=15,557), which included genotyped and non-genotyped individuals. Accordingly, the following covariates were considered: age, sex, and self-reported population groups (i.e., Native American/American Indian; Asian; Pacific Islander; African-American/Black, not of Hispanic origin; African-American/Black, of Hispanic origin; Caucasian/White, not of Hispanic origin; Caucasian/White, of Hispanic origin). In the Army STARRS cohort, the logistic regression models were applied to a fully genotyped sample of participants of European descent (N=11,235) and covariates considered were: age, sex, and the top 10 genetic PCs for population stratification adjustment.

In the Yale-Penn cohort, we conducted a multivariate GEWIS considering unrelated participants with complete genotype information (4,044 African-Americans and 3,407 European-Americans). The analysis was conducted using the recently-developed StructLMM, a linear mixed-model approach to identify and characterize loci that interact with one or more environments efficiently (18). We used the StructLMM approach to analyze whether DSM-IV criterion counts of AD, CaD, CoD, OD, and ND and the co-occurrence of multiple DSM-IV SD diagnoses interact at the same loci with respect to suicidal ideation. From the StructLMM framework, we obtained evidence of: i) loci with significant SD-related interaction effects and ii) genetic association accounting for the possibility of heterogeneous effect sizes due to multivariate SD-gene interactions. We also characterized the loci identified, estimating the fraction of genetic variance explained by multivariate SD-gene interactions and calculating the Bayes factors (BF) between the full model and models with environmental variables removed to identify which SD-related traits are most relevant for the gene interactions observed. Details regarding the statistical methods were described previously (31). The analysis was conducted separately in each major genetically-determined ancestry group (i.e., African-Americans and European-Americans). The trans-ancestry meta-analysis was conducted using METAL (33). The information regarding suicide ideation was adjusted for age, sex, genotyping array, and the top 10 PCs, and the residuals obtained were entered as phenotypic outcomes into StructLMM. To increase the discovery power of the analysis conducted in the Yale-Penn cohort, we used the single-variant association results obtained from the StructLMM approach to conduct a genome-wide gene-based analysis, using the MAGMA tool (34) implemented in the FUMA platform (35) and applying a gene-based Bonferroni multiple testing correction. Gene-based tests are more powerful than single-variant association analysis (34), because they combine single-variant signals within genic regions accounting for the linkage disequilibrium (LD) structure and reducing the multiple testing correction burden. The FUMA platform (35) was also used to perform a functional annotation of the variants identified using data from CADD (Combined Annotation Dependent Depletion) (36), RegulomeDB (37), and 15-core chromatin state information across 13 brain tissues (i.e., ganglion eminence derived primary cultured neurospheres; cortex derived primary cultured neurospheres; middle hippocampus; substantia nigra; anterior caudate; cingulate gyrus; inferior temporal lobe; angular gyrus; dorsolateral prefrontal cortex; germinal matrix; fetal female brain; fetal male brain; astrocytes). Using GTEx V8 (38), we tested the effect of the variants identified on the tissue-specific transcriptomic profiles of the surrounding genes (±1Mb of the gene transcription starting site), considering a false discovery at 5% for the genome-wide multiple testing correction. To investigate the loci identified further, we performed single-variant and gene-based phenome-wide scans leveraging more than 4,000 genome-wide summary association data collected by the GWAS Atlas (available at https://atlas.ctglab.nl/) (39).

The Army STARRS cohort was investigated to replicate the Yale-Penn findings. Due to the unavailability of information about multiple SDs among the participants, we tested the gene interaction of SUD_combined_ (i.e., a single composite variable combining alcohol and drug use disorders) with respect to SI. This analysis was conducted using the interaction test available in PLINK 1.9 (25), which compares the difference between SI regression coefficients in subjects with SUD_combined_ vs. those without SUD_combined_. Age, sex, and the top 10 PCs were entered as covariates.

A PRS analysis was conducted on Yale-Penn and Army STARRS cohorts leveraging the summary statistics of a GWAS of major depression (MD) conducted by the Psychiatric Genomics Consortium (PGC) (40). Due to the data sharing restrictions of the 23andMe personal genomics and biotechnology company (a contributor to the PGC MD cohort), GWAS data were publicly available only for a sample subset (59,851 MD cases and 113,154 controls). This analysis was restricted to the Yale-Penn and Army STARRS participants of European descent due to known biases of cross-ancestry PRS analysis (41). The MD PRS was calculated using PLINK 1.9 (25), considering multiple association P value thresholds (PT < 5×10^−8^, 10^−7^, 10^−6^, 10^−5^, 10^−4^, 0.001, 0.05, 0.1, 0.3, 0.5, 1) for SNP inclusion to identify the best-fit for each target phenotype tested. The PRS were calculated after using P-value-informed clumping with a LD cut-off of R^2^ = 0.3 within a 500kb window and excluding the major histocompatibility complex region of the genome because of its complex LD structure. The individual PRS generated were standardized and entered into a logistic regression model that included the main effect (PRS) and the effect of the interaction term (i.e., the product of PRS and the covariate for interaction).

## Results

To identify effects accounting for the comorbidity among the SDs tested in the Yale-Penn cohort, AD, CaD, CoD, ND, and OD were entered as terms in the same logistic regression model (Figure 1, left panel). We observed a consistent effect of AD and ND across all suicide traits tested, ranging from AD OR=2.11 (95%CI=1.91-2.33) and ND OR=1.42 (95%CI=1.29-1.57) for SI; to AD OR=1.66 (95%CI=1.4-1.96) and ND OR=1.29 (95%CI=1.1-1.51) for SA. Increased odds were observed with respect to CoD for SI (OR=1.69; 95%CI=1.52-1.88), SP (OR=1.26; 95%CI=1.08-1.48), and SA (OR=1.51; 95%CI=1.27-1.79), but not for persistent SI. CaD showed opposite effect directions; increased odds of SI (OR=1.62; 95%CI=1.47-1.80) and a protective effect for SA (OR=0.84; 95%CI=0.72-0.97). We observed a consistent positive relationship between the number of SD diagnoses and the odds of reporting suicidal behaviors (Figure 1, right panel). Considering the most extreme cases (i.e., all 5 SDs), we observed the largest effects: OR=6.77 (95%CI=5.74-7.99) for SI; OR=2.01 (95%CI=1.51-2.68) for persistent SI; OR=2.62 (95%CI=2.04-3.39) for SP; OR=3.61 (95%CI=2.7-4.86) for SA. In the Army STARRS participants (N=11,236), we observed similar effects of SUD_combined_: OR=2.88 (95%CI=2.6-3.19) for SI; OR=3.88 (95%CI=2.79-4.10) for SP; OR=3.92 (95%CI=3.19-4.81) for SA.

**Figure 1:**
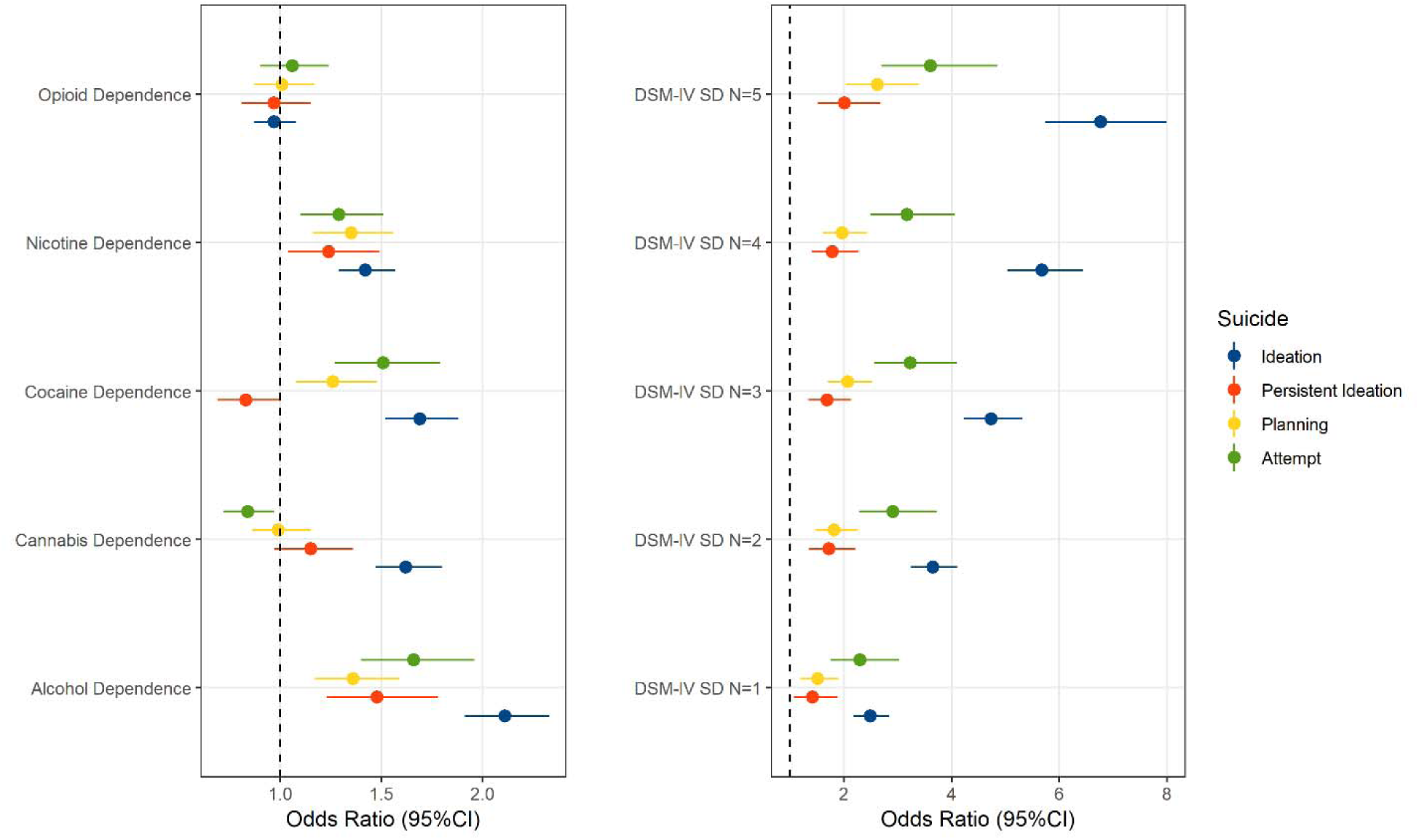
Association of DSM-IV substance dependence (SD) diagnoses (***left panel***) or polysubstance dependence severity (i.e., number of comorbid DSM-IV SD diagnosis; ***right panel***) with suicidal behaviors (ideation, persistent ideation, planning, and attempt) in Yale-Penn participants.

Among Yale-Penn participants of European descent, several genes on chromosome 16 survived multiple testing correction for both association and interactive effects (Figure 2): *LCAT* (p_association_=3.73×10^−7^; p_interaction_=1.82×10^−7^); *TSNAXIP1* (p_association_=2.08×10^−7^; p_interaction_=2.13×10^−7^), *CENPT* (p_association_=2.39×10^−7^; p_interaction_=2.32×10^−7^), and *PARD6A* (p_association_=7.17×10^−7^; p_interaction_=5.57×10^−7^). The association of this gene cluster is driven by the effect of a single variant, rs8052287 (p_association_=2.15×10^−7^; p_interaction_=7×10^−8^; Figure 3). Within this locus, 98% of the variance is explained by multivariate SD-gene interactions. We calculated the Bayes factors (BF) between the full model and models including the individual environmental exposures removed to explore which environmental variables are most relevant for the gene-environment signals of rs8052287. We observed putative gene-environment effects in rs8052287 (BF>0) for OD criterion counts (BF=12.2), CoD criterion counts (BF=12.1), ND criterion counts (BF=9.2), and co-occurrence of multiple SD diagnoses (BF=2.1). We found that rs8052287 was associated with the transcriptomic regulation of 26 genes in 30 different tissues (Supplemental Table 1). Considering brain tissues, we observed that rs8052287 is associated with *RANBP10* in the cerebellum (p=6.2×10^−18^), cortex (p=4.3×10^−12^), frontal cortex (BA9; p=4.9×10^−9^), cerebellar hemisphere (p=8.9×10^−9^), caudate (p=4.8×10^−7^), and nucleus accumbens (p=2.5×10^−5^). Rs8052287 is also a splicing quantitative trait locus for *CARMIL2* in the frontal cortex (BA9; p=1.2×10^−6^). Considering data available from the GWAS atlas, we observed 20 significant associations that survived phenome-wide multiple testing correction (Supplemental Table 2), including anthropometric traits (e.g., height p=8.30×10^−22^), male hair loss pattern (p=1.11×10^−10^), hypothyroidism (p=5.97×10^−7^), and several hematological parameters (e.g., mean corpuscular hemoglobin p=4.74×10^−7^). The same associations were also observed in the gene-based phenome-wide scans conducted for *LCAT, TSNAXIP1, CENPT*, and *PARD6A* (Supplemental Table 3). The results related to thyroid function are particularly interesting, because thyroid dysfunction has a wide range of health implications (42).

**Figure 2:**
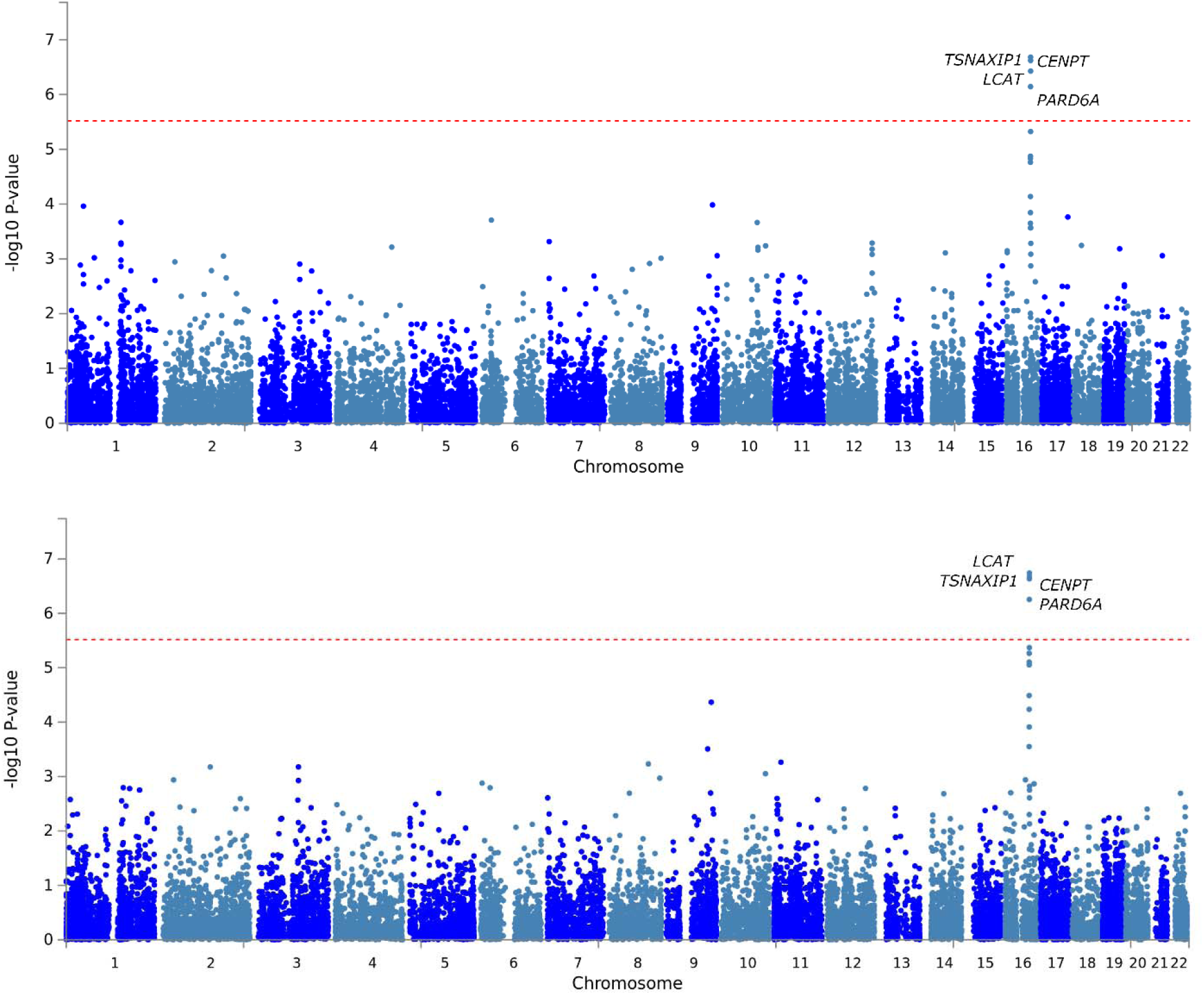
Gene-based Manhattan plots (***bottom panel***: interactive effects; ***top panel***: association effect accounting for the heterogeneous effect size due to the interactive effects) showing the results of the multivariate GEWIS of suicide ideation. Red dashed line represents the significance threshold accounting for the gene-based Bonferroni multiple testing correction.

**Figure 3:**
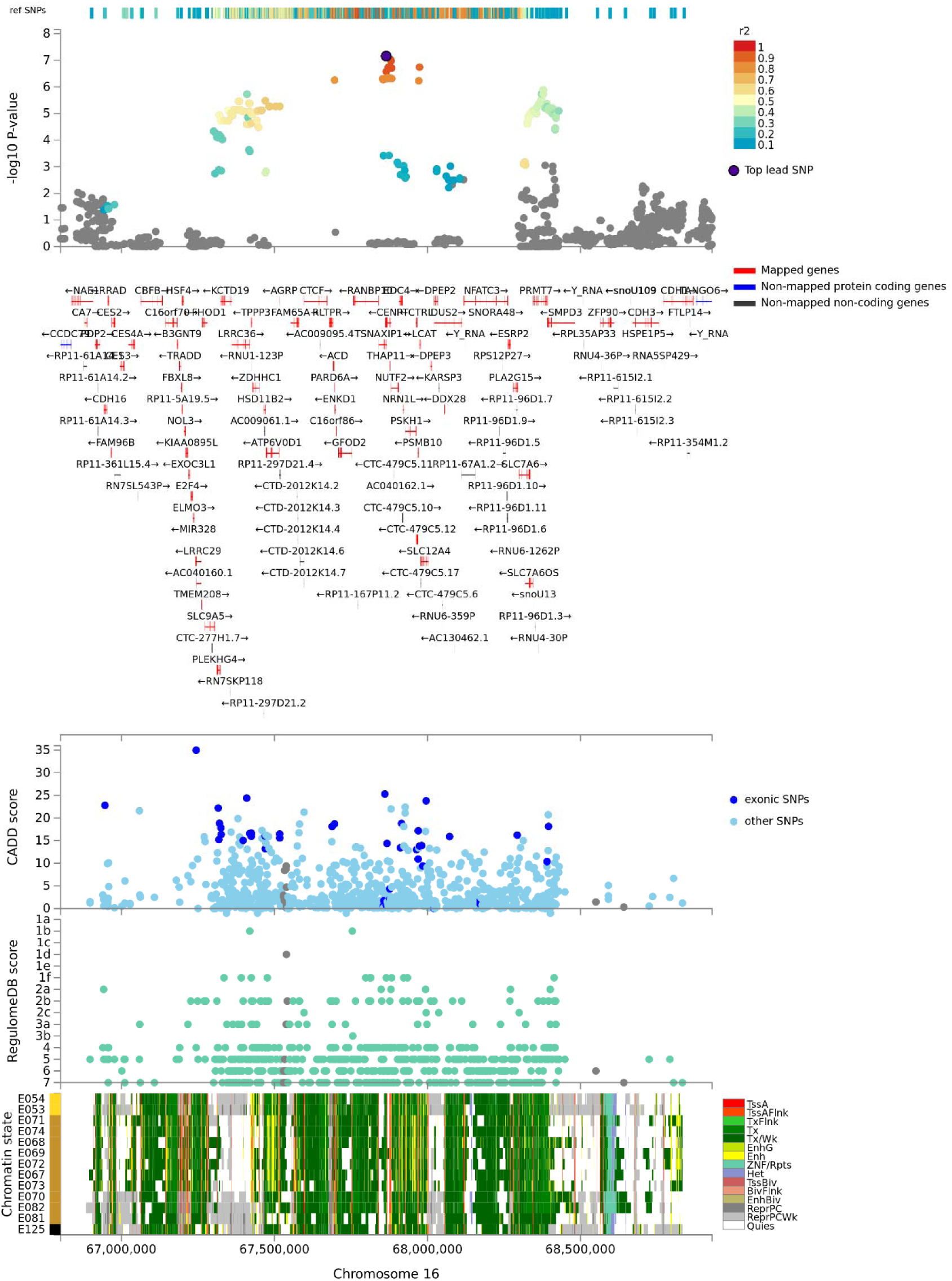
Regional Manhattan plot of the lead variant rs8052287 identified in the gene-based multivariate GEWIS of suicide ideation (Yale-Penn participants of European descent) with functional annotation derived from CADD and RegulomeDB scores and 15-core chromatin state information across 13 brain tissues.

Additionally, hypothyroidism showed genetic correlations with several behavioral traits including fatigue, anxiety, depression, loneliness, and mood swings (43). These could moderate the interplay between suicide risk and SDs. In line with the potential effect of rs8052287 on thyroid function, this variant regulates the expression of 9 genes in thyroid tissue: *LRRC36* (p=1.6×10^−16^), *ZDHHC1* (p=3.9×10^−15^), *RANBP10* (p=3.7×10^−14^), *HSD11B2* (p=5.7×10^−9^), *C16orf86* (p=3.2×10^−8^), *KCTD19* (p=8.3×10^−8^), *DUS2* (p=6.1×10^−6^), *ACD* (p=9.7×10^−5^), *FHOD1* (p=2.8×10^−4^). In addition to its effect on transcriptomic regulation, rs8052287 is in high LD (r^2^=0.81) with rs62620177, a coding variant with a CADD score of 25.3, which indicates pathogenicity in the top 1% of all SNPs in the human genome (36).

Due to the limited phenotypic information available in the Army STARRS cohort, which consisted only of a single composite SD variable (SUD_combined_), we could not apply the StructLMM approach. Testing SUD_combined_ as a single factor, no interaction of rs8052287 was observed with respect to SI outcome (p>0.05).

In the Yale-Penn participants of African descent, we observed the *HGF* gene survived Bonferroni multiple testing correction for the StructLMM interaction test (p=1.08×10^−6^; Supplemental Figure 1A) and approached significance in the association test (p=3.39×10^−6^; Supplemental Figure 1B). Within *HGF* region, we did not observe a driving single-variant association/interaction (all p>10^−5^). Although the phenome-wide scan did not show any association surviving multiple testing correction (p=1.05×10^−5^), the strongest *HGF* association was with one of the traumatic events assessed in the UK Biobank (44), Data-Field 20526: “*Been in serious accident believed to be life-threatening*” (p=1.21×10^−5^).

Based on the recent GWAS of suicidal behaviors (9-13), we focused our attention on MD PRS from a large-scale MD GWAS conducted by PGC investigators (40). In both the Yale-Penn and Army STARRS cohorts, the MD PRS was associated with increased odds of suicidal behaviors (Supplemental Table 4): SI (Yale-Penn OR=1.10, 95%CI=1-1.21; Army STARRS OR=1.09, 95%CI=1.03-1.15), persistent SI (Yale-Penn OR=1.26, 95%CI=1.09-1.46), SP (Yale-Penn OR=1.28, 95%CI=1.13-1.47; Army STARRS OR=1.07, 95%CI=0.97-1.18), and SA (Yale-Penn OR=1.26, 95%CI=1.09-1.45; Army STARRS OR=1.11, 95%CI=1-1.23). However, there was no interaction between MD PRS and SD-related traits with respect to the suicidal behaviors tested (Supplemental Table 5).

## Discussion

To our knowledge, this is the first study to investigate polysubstance dependence in relation to the spectrum of suicidal behaviors and to use high-dimensional data to identify multi-environment gene interactions related to the interplay of these complex phenotypes.

Leveraging the deep phenotypic assessment available in the Yale-Penn cohort, we investigated the effects of different SDs on the odds of suicidal behaviors. Each SD appears to have a specific pattern of suicide-related associations. AD and ND consistently increased the odds of all suicidal behaviors tested (i.e., ideation, persistent ideation, planning, and attempt). The findings are in line with previous studies and meta-analyses (3, 45). In contrast, the other SDs investigated appear not to have the same effect across suicidal behaviors but rather specifically affect certain suicidal states. The most intriguing results are related to CaD, which was associated with increased odds of SI, a null effect on persistent SI and SP, and an association with reduced odds of SA. A meta-analysis of studies of on cannabis use and suicidality showed a positive association (i.e., increased risk) of cannabis use on risk of SI, SA, and death by suicide (46). However, this study recognized a lack of homogeneity in the measurement of cannabis exposure and systematic control for potential confounders as key limitations. Our analysis was based on a comprehensive assessment of multiple SDs including CaD, and, for that specific trait, the results are controlled for the effect of other SDs on suicidal behaviors. These methodological differences could explain the inconsistency between our findings and those of the previous meta-analysis. Further, the previous meta-analysis focused on varying degrees of cannabis use (any, chronic, and heavy cannabis use), while our analysis investigated CaD. Recent studies have shown that the genetic basis of substance use (in terms of quantity and frequency used) and SD (in terms of physiological dependence) are partially distinct (47, 48) and these differences may affect their associations with suicidal behaviors.

Beyond the effect of single SDs, we tested whether the severity of polysubstance dependence (i.e., the number of SD diagnoses among Yale-Penn participants) affects the odds of suicidal behaviors. Previous studies generally investigated polysubstance dependence as a binary outcome, considering individuals with two or more SDs as cases (49-51). Using quantitative approach, we observed that individuals with increasing number of SD diagnoses have increased odds of suicidal behaviors.

Interestingly, this effect showed a non-linear trend across the severity of suicide behaviors, where the strongest associations were observed for SI (individuals with all five SD diagnoses showed a 6.77-fold increase in SI odds) and SA (individuals with all five SD diagnoses showed a 3.61-fold increase in SA odds) and relatively weaker associations were present for persistent SI (individuals with all five SD diagnoses showed a 2.01-fold increase in persistent-SI odds) and SP (individuals with all five SD diagnoses showed a 2.62-fold increase in SP odds). Thus, the relationship between SDs and suicidal behaviors does not follow the escalation of suicidality along a continuum in subjects affected by polysubstance dependence. However, our analysis in the Army STARRS participants showed a linear trend where individuals with SUD_combined_ have odds that increase with the seriousness of suicidal behaviors (SI OR=2.88; SP OR=3.38; SA OR=3.92). The difference between Yale-Penn and Army STARRS results may be due to the analytic designs, in that we could not calculate the severity of polysubstance dependence because information regarding different SD diagnoses was not available in the Army STARRS assessment. However, the different phenotypic characteristics of Yale-Penn and Army STARRS cohorts may also have contributed to the different outcomes observed.

We conducted a multivariate gene-based GEWIS of SI, testing the interactive effect of SD-related traits. In participants of European descent, we identified multiple genes within the same region of chromosome 16 that showed both significant SD-related interaction effects and a significant association with SI accounting for the possibility of heterogeneous effect sizes due to multivariate SD-gene interactions. The analysis of the single-variant effects within this chromosomal region showed that the association is partially attributable to a single variant. Investigating the index variant, rs8052287, we characterized the signal and observed that the multivariate interactions were related to DSM-IV OD, CoD, and ND criterion counts and the severity of polysubstance dependence (i.e., the number of SD diagnoses). These gene-environment interactions account for 98% of the genetic variance within this locus. This is in line with the expected statistical power of the StructLMM method, which has greater power to detect loci with a high fraction of the genetic variance explained by gene-environment interactions (18). We could not apply the StructLMM method to the Army STARRS data because of the lack of high-dimensional data on polysubstance dependence. Applying a standard gene-environment test, we did not observe an interaction between rs8052287 and SUD_combined_ with respect to SI. This is probably due to the dramatic reduction in the statistical power of standard gene-environment tests compared to the StructLMM approach (18).

To validate our findings, we investigated the phenome-wide spectrum associated with this locus and the regulatory effect of rs8052287 (i.e., the index variant) on the tissue-specific transcriptomic profile of the genes located in this region. Gene-based and single-variant phenome-wide scans showed a similar pattern of associations related to physical health and characteristics. These included hypothyroidism, anthropometric traits, male hair loss, and hematological parameters. These phenotypic associations can be linked to altered thyroid function (42). Tissue-specific transcriptomic analysis confirmed that rs8052287 actively regulates multiple genes in thyroid tissues. Due to the high gene density in the locus identified (rs8052287 regulates the transcriptomic profile of 26 genes), it is hard to pinpoint the gene(s) responsible for the interaction with polysubstance dependence. However, the evidence leading to altered thyroid function supports an intriguing hypothesis. Previous studies hypothesized a role of thyroid dysfunction in suicide risk, especially among psychiatric patients (52-54). This is in line with the known effect of altered thyroid function on mental health (42). In a recent genome-wide analysis, we observed that hypothyroidism is genetically correlated with several behavioral traits including fatigue, anxiety, depression, loneliness, and mood swings (43). Based on these findings, we hypothesize that the region identified may affect suicide risk via its role in regulating thyroid function. Additionally, the three substances (cocaine, nicotine, and opioids) that showed an interactive effect with rs8052287 on SI have been investigated previously with respect to thyroid homeostasis. Cigarette smoking appears to affect thyroid function with a dose-related effect linking cotinine levels (a nicotine metabolite (55)) to thyroid function and thyroid autoimmunity (56). Cocaine abuse has been linked to the disruptions in the hypothalamic-pituitary-thyroid axis (57) and cocaine use has been suggested as a possible trigger for thyroid storm (58). There is a growing literature on the effect of opioids on the endocrine system and opioid-induced endocrinopathies (59). Acute administration of various opioids has been shown to alter thyroid-stimulating hormone and thyrotropin-releasing hormone consistenly (59). However, conflicting results were obtained by studies investigating thyroid function in long-term opioid users and controls (59). Although the exact pathway by which the combination of CoD, ND, and OD interacts with rs8052287 in increasing SI risk is unclear, our “thyroid” hypothesis provides a potential pathogenetic mechanism linking polysubstance dependence, genetic liability to hypothyroidism, and SI. Further studies are needed to test this hypothesis, which has some obvious potential therapeutic implications.

In conclusion, we provide novel insights into the epidemiology of suicidal behaviors and polysubstance dependence, identifying the effects of five SDs while controlling for their comorbidity and assessing their combined effect. Our multivariate GEWIS of SI identified a locus with supporting phenomic and transcriptomic evidences. To replicate these findings, future studies will require cohorts with the same high-dimensional data available in the Yale-Penn sample. Finally, the highly polygenic nature of psychiatric traits will require a much larger sample size to identify a representative fraction of the genetic predisposition to suicidal behaviors in the context of SDs.

## Data Availability

Data generated are included in the manuscript and the supplemental material.

## Acknowledgements

This study was supported by the American Foundation for Suicide Prevention (YIG-1-109-16), the National Institute on Drug Abuse (R21 DA047527), and the Veterans Affairs National Center for Posttraumatic Stress Disorder Research. The Yale-Penn cohort was supported by multiple grants from the National Institutes of Health (R01 DA12690, R01 AA11330, and R01 AA017535). Army STARRS was sponsored by the Department of the Army and funded under U01MH087981 (2009-2015) with NIMH.

## References

1. Yuodelis-Flores C., Ries R. K. Addiction and suicide: A review, Am J Addict 2015: 24: 98–104.

2. Roy A. Risk factors for attempting suicide in heroin addicts, Suicide Life Threat Behav 2010: 40: 416–420.

3. Pompili M., Serafini G., Innamorati M., Dominici G., Ferracuti S., Kotzalidis G. D. et al. Suicidal behavior and alcohol abuse, Int J Environ Res Public Health 2010: 7: 1392–1431.

4. Roy A. Characteristics of cocaine dependent patients who attempt suicide, Arch Suicide Res 2009: 13: 46–51.

5. Zai C. C., De Luca V., Strauss J., Tong R. P., Sakinofsky I., Kennedy J. L. Genetic Factors and Suicidal Behavior. In: Dwivedi Y., editor. The Neurobiological Basis of Suicide, Boca Raton (FL); 2012.

6. Mullins N., Perroud N., Uher R., Butler A. W., Cohen-Woods S., Rivera M. et al. Genetic relationships between suicide attempts, suicidal ideation and major psychiatric disorders: a genome-wide association and polygenic scoring study, Am J Med Genet B Neuropsychiatr Genet 2014: 165B: 428–437.

7. Border R., Johnson E. C., Evans L. M., Smolen A., Berley N., Sullivan P. F. et al. No Support for Historical Candidate Gene or Candidate Gene-by-Interaction Hypotheses for Major Depression Across Multiple Large Samples, Am J Psychiatry 2019: 176: 376–387.

8. Johnson E. C., Border R., Melroy-Greif W. E., De Leeuw C. A., Ehringer M. A., Keller M. C. No Evidence That Schizophrenia Candidate Genes Are More Associated With Schizophrenia Than Noncandidate Genes, Biol Psychiatry 2017: 82: 702–708.

9. Mullins N., Bigdeli T. B., Borglum A. D., Coleman J. R. I., Demontis D., Mehta D. et al. GWAS of Suicide Attempt in Psychiatric Disorders and Association With Major Depression Polygenic Risk Scores, Am J Psychiatry 2019: 176: 651–660.

10. Stein M. B., Ware E. B., Mitchell C., Chen C. Y., Borja S., Cai T. et al. Genomewide association studies of suicide attempts in US soldiers, Am J Med Genet B Neuropsychiatr Genet 2017: 174: 786–797.

11. Otsuka I., Akiyama M., Shirakawa O., Okazaki S., Momozawa Y., Kamatani Y. et al. Genome-wide association studies identify polygenic effects for completed suicide in the Japanese population, Neuropsychopharmacology 2019: 44: 2119–2124.

12. Strawbridge R. J., Ward J., Ferguson A., Graham N., Shaw R. J., Cullen B. et al. Identification of novel genome-wide associations for suicidality in UK Biobank, genetic correlation with psychiatric disorders and polygenic association with completed suicide, EBioMedicine 2019: 41: 517–525.

13. Levey D. F., Polimanti R., Cheng Z., Zhou H., Nunez Y. Z., Jain S. et al. Genetic associations with suicide attempt severity and genetic overlap with major depression, Transl Psychiatry 2019: 9: 22.

14. Polimanti R., Wang Q., Meda S. A., Patel K. T., Pearlson G. D., Zhao H. et al. The Interplay Between Risky Sexual Behaviors and Alcohol Dependence: Genome-Wide Association and Neuroimaging Support for LHPP as a Risk Gene, Neuropsychopharmacology 2017: 42: 598–605.

15. Polimanti R., Kaufman J., Zhao H., Kranzler H. R., Ursano R. J., Kessler R. C. et al. A genome-wide gene-by-trauma interaction study of alcohol misuse in two independent cohorts identifies PRKG1 as a risk locus, Mol Psychiatry 2018: 23: 154–160.

16. Polimanti R., Meda S. A., Pearlson G. D., Zhao H., Sherva R., Farrer L. A. et al. S100A10 identified in a genome-wide gene x cannabis dependence interaction analysis of risky sexual behaviours, J Psychiatry Neurosci 2017: 42: 252–261.

17. Polimanti R., Zhao H., Farrer L. A., Kranzler H. R., Gelernter J. Ancestry-specific and sex-specific risk alleles identified in a genome-wide gene-by-alcohol dependence interaction study of risky sexual behaviors, Am J Med Genet B Neuropsychiatr Genet 2017: 174: 846–853.

18. Moore R., Casale F. P., Jan Bonder M., Horta D., Consortium B., Franke L. et al. A linear mixed-model approach to study multivariate gene-environment interactions, Nat Genet 2019: 51: 180–186.

19. Gelernter J., Sherva R., Koesterer R., Almasy L., Zhao H., Kranzler H. R. et al. Genome-wide association study of cocaine dependence and related traits: FAM53B identified as a risk gene, Mol Psychiatry 2014: 19: 717–723.

20. Gelernter J., Kranzler H. R., Sherva R., Almasy L., Herman A. I., Koesterer R. et al. Genome-wide association study of nicotine dependence in American populations: identification of novel risk loci in both African-Americans and European-Americans, Biol Psychiatry 2015: 77: 493–503.

21. Gelernter J., Kranzler H. R., Sherva R., Almasy L., Koesterer R., Smith A. H. et al. Genome-wide association study of alcohol dependence:significant findings in African- and European-Americans including novel risk loci, Mol Psychiatry 2014: 19: 41–49.

22. Sherva R., Wang Q., Kranzler H., Zhao H., Koesterer R., Herman A. et al. Genome-wide Association Study of Cannabis Dependence Severity, Novel Risk Variants, and Shared Genetic Risks, JAMA Psychiatry 2016: 73: 472–480.

23. Gelernter J., Kranzler H. R., Sherva R., Koesterer R., Almasy L., Zhao H. et al. Genome-wide association study of opioid dependence: multiple associations mapped to calcium and potassium pathways, Biol Psychiatry 2014: 76: 66–74.

24. Pierucci-Lagha A., Gelernter J., Feinn R., Cubells J. F., Pearson D., Pollastri A. et al. Diagnostic reliability of the Semi-structured Assessment for Drug Dependence and Alcoholism (SSADDA), Drug Alcohol Depend 2005: 80: 303–312.

25. Chang C. C., Chow C. C., Tellier L. C., Vattikuti S., Purcell S. M., Lee J. J. Second-generation PLINK: rising to the challenge of larger and richer datasets, Gigascience 2015: 4: 7.

26. Walters R. K., Polimanti R., Johnson E. C., Mcclintick J. N., Adams M. J., Adkins A. E. et al. Transancestral GWAS of alcohol dependence reveals common genetic underpinnings with psychiatric disorders, Nat Neurosci 2018: 21: 1656–1669.

27. Ursano R. J., Colpe L. J., Heeringa S. G., Kessler R. C., Schoenbaum M., Stein M. B. et al. The Army study to assess risk and resilience in servicemembers (Army STARRS), Psychiatry 2014: 77: 107–119.

28. Kessler R. C., Ustun T. B. The World Mental Health (WMH) Survey Initiative Version of the World Health Organization (WHO) Composite International Diagnostic Interview (CIDI), Int J Methods Psychiatr Res 2004: 13: 93–121.

29. Nock M. K., Stein M. B., Heeringa S. G., Ursano R. J., Colpe L. J., Fullerton C. S. et al. Prevalence and correlates of suicidal behavior among soldiers: results from the Army Study to Assess Risk and Resilience in Servicemembers (Army STARRS), JAMA Psychiatry 2014: 71: 514–522.

30. Posner K., Brown G. K., Stanley B., Brent D. A., Yershova K. V., Oquendo M. et al. The Columbia-Suicide Severity Rating Scale: initial validity and internal consistency findings from three multisite studies with adolescents and adults, Am J Psychiatry 2011: 168: 1266–1277.

31. Stein M. B., Chen C. Y., Ursano R. J., Cai T., Gelernter J., Heeringa S. G. et al. Genome-wide Association Studies of Posttraumatic Stress Disorder in 2 Cohorts of US Army Soldiers, JAMA Psychiatry 2016: 73: 695–704.

32. R CORE TEAM. R: A language and environment for statistical computing. R Foundation for Statistical Computing, Vienna, Austria; 2014.

33. Willer C. J., Li Y., Abecasis G. R. METAL: fast and efficient meta-analysis of genomewide association scans, Bioinformatics 2010: 26: 2190–2191.

34. De Leeuw C. A., Mooij J. M., Heskes T., Posthuma D. MAGMA: generalized gene-set analysis of GWAS data, PLoS Comput Biol 2015: 11: e1004219.

35. Watanabe K., Taskesen E., Van Bochoven A., Posthuma D. Functional mapping and annotation of genetic associations with FUMA, Nat Commun 2017: 8: 1826.

36. Rentzsch P., Witten D., Cooper G. M., Shendure J., Kircher M. CADD: predicting the deleteriousness of variants throughout the human genome, Nucleic Acids Res 2019: 47: D886–D894.

37. Boyle A. P., Hong E. L., Hariharan M., Cheng Y., Schaub M. A., Kasowski M. et al. Annotation of functional variation in personal genomes using RegulomeDB, Genome Res 2012: 22: 1790–1797.

38. Consortium G. T., Laboratory D. A., COORDINATING CENTER -ANALYSIS WORKING G., STATISTICAL METHODS GROUPS-ANALYSIS WORKING G., Enhancing G. G., Fund N. I. H. C. et al. Genetic effects on gene expression across human tissues, Nature 2017: 550: 204–213.

39. Watanabe K., Stringer S., Frei O., Umicevic Mirkov M., De Leeuw C., Polderman T. J. C. et al. A global overview of pleiotropy and genetic architecture in complex traits, Nat Genet 2019: 51: 1339–1348.

40. Wray N. R., Ripke S., Mattheisen M., Trzaskowski M., Byrne E. M. ABDELLAOUI et al. Genome-wide association analyses identify 44 risk variants and refine the genetic architecture of major depression, Nat Genet 2018: 50: 668–681.

41. Martin A. R., Gignoux C. R., Walters R. K., Wojcik G. L., Neale B. M., Gravel S. et al. Human Demographic History Impacts Genetic Risk Prediction across Diverse Populations, Am J Hum Genet 2017: 100: 635–649.

42. Taylor P. N., Albrecht D., Scholz A., Gutierrez-Buey G., Lazarus J. H., Dayan C. M. et al. Global epidemiology of hyperthyroidism and hypothyroidism, Nat Rev Endocrinol 2018: 14: 301–316.

43. Ravera S., Carrasco N., Gelernter J., Polimanti R. Phenomic Impact of Genetically-Determined Euthyroid Function and Molecular Differences between Thyroid Disorders, J Clin Med 2018: 7.

44. Bycroft C., Freeman C., Petkova D., Band G., Elliott L. T., Sharp K. et al. The UK Biobank resource with deep phenotyping and genomic data, Nature 2018: 562: 203–209.

45. Hughes J. R. Smoking and suicide: a brief overview, Drug Alcohol Depend 2008: 98: 169–178.

46. Borges G., Bagge C. L., Orozco R. A literature review and meta-analyses of cannabis use and suicidality, J Affect Disord 2016: 195: 63–74.

47. Kranzler H. R., Zhou H., Kember R. L., Vickers Smith R., Justice A. C., Damrauer S. et al. Genome-wide association study of alcohol consumption and use disorder in 274,424 individuals from multiple populations, Nat Commun 2019: 10: 1499.

48. Polimanti R., Walters R. K., Johnson E. C., Mcclintick J. N., Adkins A. E., Adkins D. E. et al. Leveraging genome-wide data to investigate differences between opioid use <em>vs</em>. opioid dependence in 41,176 individuals from the Psychiatric Genomics Consortium, bioRxiv 2019: 765065.

49. Martinotti G., Carli V., Tedeschi D., Di Giannantonio M., Roy A., Janiri L. et al. Mono- and polysubstance dependent subjects differ on social factors, childhood trauma, personality, suicidal behaviour, and comorbid Axis I diagnoses, Addict Behav 2009: 34: 790–793.

50. Verdoux H., Liraud F., Gonzales B., Assens F., Abalan F., Van Os J. Suicidality and substance misuse in first-admitted subjects with psychotic disorder, Acta Psychiatr Scand 1999: 100: 389–395.

51. Guvendeger Doksat N., Zahmacioglu O., Ciftci Demirci A., Kocaman G. M., Erdogan A. Association of Suicide Attempts and Non-Suicidal Self-Injury Behaviors With Substance Use and Family Characteristics Among Children and Adolescents Seeking Treatment for Substance Use Disorder, Subst Use Misuse 2017: 52: 604–613.

52. Shen Y., Wu F., Zhou Y., Ma Y., Huang X., Ning Y. et al. Association of thyroid dysfunction with suicide attempts in first-episode and drug naive patients with major depressive disorder, J Affect Disord 2019: 259: 180–185.

53. Charlier P., Watier L., Menetrier M., Chaillot P. F., Brun L., De La Grandmaison G. L. Is suicide risk correlated to thyroid weight?, Med Hypotheses 2012: 79: 264–266.

54. Jose J., Nandeesha H., Kattimani S., Meiyappan K., Sarkar S., Sivasankar D. Association between prolactin and thyroid hormones with severity of psychopathology and suicide risk in drug free male schizophrenia, Clin Chim Acta 2015: 444: 78–80.

55. Benowitz N. L., Pomerleau O. F., Pomerleau C. S., Jacob P., 3rd. Nicotine metabolite ratio as a predictor of cigarette consumption, Nicotine Tob Res 2003: 5: 621–624.

56. Kim S. J., Kim M. J., Yoon S. G., Myong J. P., Yu H. W., Chai Y. J. et al. Impact of smoking on thyroid gland: dose-related effect of urinary cotinine levels on thyroid function and thyroid autoimmunity, Sci Rep 2019: 9: 4213.

57. Kovalevich J., Corley G., Yen W., Kim J., Rawls S. M., Langford D. Cocaine decreases expression of neurogranin via alterations in thyroid receptor/retinoid X receptor signaling, J Neurochem 2012: 121: 302–313.

58. Lacy M. E., Utzschneider K. M. Cocaine Intoxication and Thyroid Storm: Similarity in Presentation and Implications for Treatment, J Investig Med High Impact Case Rep 2014: 2: 2324709614554836.

59. Fountas A., Van Uum S., Karavitaki N. Opioid-induced endocrinopathies, Lancet Diabetes Endocrinol 2019.

